# Hardware-Powered Ultra Low Latency (HarPULL) Brain State Dependent TMS Technology

**DOI:** 10.1101/2024.05.03.24306813

**Authors:** Milana Makarova, Nikita Fedosov, Julia Nekrasova, Alexey Ossadtchi

**Affiliations:** Center for Bioelectric Interfaces, Higher School of Economics, Moscow, Russia; AIRI, Artificial Intelligence Research Institute, Moscow, Russia; LLC “Life Improvement by Future Technologies Center”, Moscow, Russia

## Abstract

**Objective:** Upcoming neuroscientific research will require bidirectional and context dependent interaction with nervous tissue. To facilitate the future neuroscientific discoveries we have created HarPULL, a genuinely real-time system for tracking oscillatory brain state.

**Approach:** The HarPULL technology ensures reliable, accurate and affordable real-time phase and amplitude tracking based on the state-space estimation framework operationalized by Kalman filtering. To avoid data transfer delays and to obtain a truly real-time system the algorithm is implemented on the computational core of an EEG amplifier controlled by a real-time operating system. Systems performance is tested with simulated and real data both online and offline and within a real-time state dependent TMS using a phantom and human subjects.

**Main results:** We show that taking into account the 1*/f* nature of the brain noise and the use of the steady state colored Kalman filter further improves phase tracking performance in both simulated and real data. We use HarPULL to trigger the TMS device contingent upon the target phase and amplitude combination and demonstrate minimal delay (2 ms) between the occurrence of the predetermined rhythm phase in the cortex and the corresponding magnetic stimulus. Using this setup in the real-time setting we observe a significant modulation of the motor evoked potentials (MEP) by the sensorimotor rhythm’s state. Finally, we use HarPULL and for the first time obtain phase-dependent muscle cortical representation (MCR) maps in real-time. We show better delineation between the representations of several muscles when the stimulation is performed in the excitation state.

**Significance:** HarPULL is the first truly real-time technology for the instantaneous tracking of the brain’s rhythmic activity. Our technological solution establishes a nearly instantaneous non-invasive contact with a living brain which has a broad range of clinical, diagnostic and scientific applications.

## 1 Introduction

Transcranial magnetic stimulation (TMS) generally provides effective, high-resolution (on the level of millimeters and milliseconds), yet non-invasive and safe interaction with neuronal ensembles of the brain [7]. Combining TMS with electromyography (EMG), electroencephalography (EEG), functional near infra-red spectroscopy (fNIRS), transcranial electric stimulation (TES: tACS and tDCS) techniques and even functional magnetic resonance imaging (fMRI) allows us to quantify and modulate neuronal activity and measure the stimulation effect immediately or in the subsequent time periods. These opportunities hold great promise in diagnostics and prognostics of neurological and psychiatric disorders, therapy and rehabilitation, functional mapping and assessment of cortical excitability [29, 58]. However, unlike the peripheral nerve fibers whose stimulation leads to stable and easily interpretable responses in the target muscle, stimulation of the conductive pathways of the central nervous system (CNS) does not furnish persistent and reproducible reactions. Excessive trial-to-trial and long-term amplitude variability of the evoked brain and cortico-spinal responses (also known as motor evoked potentials - MEPs) both within and across subjects considerably reduces the enthusiasm in the community towards this potentially exciting technology [18, 31, 37].

The reason for this obstacle is the complexity, both anatomical and functional, of the CNS tracts and their modulation by a range of functional systems of the brain so that the current reaction of an organism to the presented stimulus depends on the current brain state [49]. While “brain state” is somewhat hard to define, to some extent and in the context of the specific paradigm it can be associated with the phase and amplitude of the spontaneous oscillations (brain rhythms). This way “brain state” derives from the periodical succession of the excitation and inhibition intervals of the neuronal populations caused by synchronized changes in the membrane potentials of neuronal populations [8, 10, 44].

To date, there exists a large body of evidence regarding the specific relations between the executive and motor functions and instantaneous phase and/or amplitude of slow-wave rhythms (most commonly in alpha - both occipital and sensorimotor, or theta-range). This has been confirmed in animal recordings *in vivo* [1, 4, *17], in vitro* [21, 22] and human non-invasive recordings (including the use of TMS) [2, 3, 6, 9, 14, 16, 19, 23, 24, 26, 30, 32, 34, 40, 41, 45, 47, 57, 59, 64, 66]. Despite the fact that the questions about the strength of this correlational relationship and its direction remain open, see [17, 45, 64] vs. [6, 23, 24, 26, 32, 47, 56, 59], it is conceivable that cortical rhythms significantly modulate both neuronal and behavioral responses and therefore can be considered as encoders of the brain state.

It also remains unclear which of the morphological characteristics of the rhythmic brain activity (i.e., power, phase, burst incidence rate) or their combination would be the most appropriate control signal for effective state-dependent interventions [27]. The instantaneous phase value in theory represents a measure of neuronal ensemble excitability and appears to be a stable predictor of higher MEP amplitudes at the trough relative to the peak of sensorimotor rhythm [46, 55, 66]. Rhythm’s amplitude, on the other hand, was shown to modulate the normalized firing rate in the sensorimotor regions of monkeys, with an inverse relationship with alpha power [17]. In other studies, a similar correlation with alpha power was either not found at all [6, 23, 24, 26, 32, 47, 59] or was reversed [56]. The overall complex nature of EEG time series and the presence of oscillations in various frequency bands in particular requires application of signal processing algorithms to extract the parameters of rhythmic activity within the band of interest. Viewing macroscopically observed brain rhythm as a frequency modulated process [62] we can conclude that the fluctuations of instantaneous phase is driven by the continuously changing instantaneous frequency. Gabor’s uncertainty principle limits the achievable combination of temporal and frequency resolution which requires finding signal processing methods that furnish an acceptable trade-off between phase estimation accuracy and the incurred delay.

There is a number of algorithms aimed at improving the accuracy and minimizing delays [12, 33, 43, 50, 51, 63]. Most of them can be divided into two major groups: AR-based techniques and dynamic model based approaches, operationalized by Kalman filtering. AR-based models rely on windowed techniques (like band-pass filtering followed by the Hilbert transform and phase estimation). Both of these operations introduce fundamental delays and forward-prediction methods such as autoregressive (AR) models or linear phase extrapolation are used to compensate for it. Broadband and often non-sinusoidal EEG signal with occasional phase resets leaves little opportunity to develop an accurate filter, which does not significantly distort the waveform, and not to misinterpret the phase [11, 13, 63]. The AR-based techniques incur additional computations, which may be a burden in a trully real-time setting [35], and also do not account for measurement noise and a generative model of the signal. Some of the algorithms are available as toolboxes, like PHASTIMATE [67] or SSPE method from Open Ephys [63].

Moreover, in addition to the algorithmic delays required for phase estimation, practical PC-based applications of the approaches described above incurs also purely technical delays. They are associated with data transfer between the EEG acquisition device, computing and feedback presentation units (e.g. TMS stimulator). This additional latency is influenced by the hard to control processes on-going in the personal computer typically run under the non-real-time operational system. [60].

Thus, there is a demand to develop a technology for close-to-instantaneous accurate tracking of brain rhythm parameters capable of delivering stimuli associated with particular neuronal states defined by the combination of rhythm’s instantaneous phase and amplitude. This would allow not only to achieve the improved accuracy and reproducibility of TMS stimulation studies, but also to significantly reduce the time of the experiment aimed at both research and diagnostics of the cortico-spinal responses. Combined with and enabling novel real-time experimental paradigms such a technology would help to unravel the complex causes of MEP generation mechanisms. More globally, this technology would not be limited to controlling the TMS as a feedback device but would rather become instrumental in establishing reliable and informative, dynamic and interactive contact with the nervous tissue in both non-invasive and invasive settings. Combined with and enabling novel real-time experimental paradigms such a technology will help to unravel the complex causes of mechanisms of MEP fluctuation and unravelling

To this end, we have developed a hardware-powered ultra-low latency (HarPULL) system for robust real-time tracking of brain rhythms’ phase and envelope. Our setup is portable, allows for generation of real-time low latency stimuli contingent upon user specified combination of the target brain rhythm parameters and runs on-board of a regular reprogrammed EEG device. Our solution offers:

1. accurate, low latency instantaneous phase and envelope estimates achieved by a computationally light and robust algorithm for decomposing EEG time series into several rhythmic components modelled as a frequency modulated process [35];
2. regular PC application the workflow control that performs short segment of training data acquisition, rhythm’s model parameters identification and their transfer on board to the reprogrammed EEG device hardware for real-time processing and oscillatory state detection;
3. hardware implementation on board of a regular EEG acquisition device which furnishes 2 ms real-time delay between the moment of occurrence of the desired event (the phase of the tracked rhythm) in the brain activity and the arrival of the trigger stimulus used to release TMS stimulation pulse or initiate other forms of neurofeedback.

In what follows after outlining the methodological components of our solution, we proceed to validate its algorithmic aspects using a combination of simulated and real data, including the datasets provided by Zrenner et al. [67]. We then describe our further experiments designed to confirm the overall HarPULL’s functionality, specifically its low-latency phase estimation property. To this end we performed a closed-loop TMS experiment using ripe watermelon as a naturalistic model whose electrical activity was driven by an Arduino-controlled current driver.

Next, we detail actual human subjects experiments utilizing our HarPULL system and demonstrating a clearly pronounced modulation of MEP amplitude by the ongoing sensorimotor rhythm phase at stimulation moment. Additionally, we present muscle cortical representation maps obtained for the first time using real-time state dependent TMS using two fixed target phase values whose acquisition required only a fraction of time and TMS pulses as compared to the more traditional phase agnostic setting. In conclusion, we summarize our solution, discuss the results and outline its potential uses, future research vista and the avenues for further development.

## 2 Materials and Methods

### 2.1 State-space modeling framework

In order to estimate phase and envelope in real-time and with minimal latency we rely on a linear Matsuda-Komaki model [35] providing a decomposition of EEG time series into a superposition of rhythmic components. This approach uses Kalman filtering (KF) to track the states of rhythmic components each modeled as a frequency modulated process with its own central frequency and depth of stochastic modulation. The idea of modeling brain rhythms as frequency modulated processes dates back to N. Wiener who suggested it in 1960, see [62] for a review. Based on the recent reports the state-space modeling approach offers significant advantages [63] as compared to the competing methods. The technique relies on two separate equations: the recurrent state evolution equation, which reflects the underlying dynamics of the target process (brain rhythm) and the observation equation, which transforms the state into the observed signal. In order to facilitate subsequent estimation of the instantaneous phase and amplitude of the ongoing brain rhythm each oscillation is described by a state vector with two entries ***x***_*t*_ = [*x*_*t*,1_, *x*_*t*,2_]^*T*^ corresponding to the real and imaginary parts of the associated analytic signal *z*_*t*_ = *x*_*t*,1_ + *jx*_*t*,2_, whose vector rotates in the complex plane with mean angular velocity 2*πf*_0_ where *f*_0_ is the rhythm’s central frequency. Then, a single oscillation time series is described by the following recursive equation [35] :

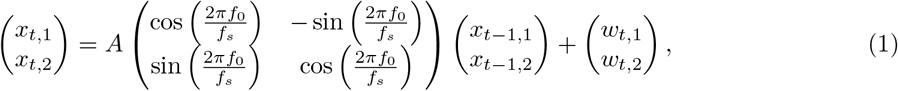

where

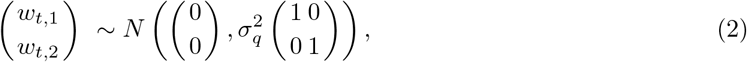

Here *f*_*s*_ is the sampling frequency of our data so that dimensionless digital frequency 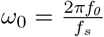 is always between 0 and *π*, 0 *< A <* 1 - is a damping coefficient which makes the model stable and defines the band-width of the resultant frequency modulated process. The model is driven by the vector of temporally white Gaussian noise ***w***_*t*_ = [*w*_*t*,1_, *w*_*t*,2_]^*T*^ with diagonal covariance 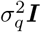.

The observed scalar sequence *y*_*t*_ is then simply an instantaneous mixture of the first state vector element corresponding to the real-part of the rhythm’s analytic signal and samples *v*_*t*_ of Gaussian noise with variance 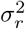:

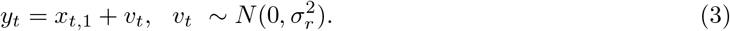

In the classical model described by [35] *v*_*t*_ is assumed to be temporally white. Later here we also demonstrate a solution where we extend this model to capture the 1*/f* nature of brain noise using a high order auto-regressive model to capture background brain activity in the close to DC frequency range. Defining rhythm’s evolution matrix 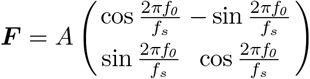 and observations matrix ***H*** = **h**^*T*^ = [1, 0] we can write these equations in the form more conventional for subsequent Kalman filtering [25] based inference of the oscillatory state vector ***x***_*t*_ from the observations *y*_*t*_ under the assumption of temporally white observation noise *v*_*t*_.

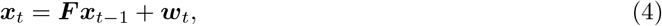

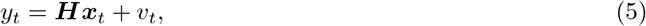

Using conventional notation for the driving noise covariance ***Q***_*t*_ = *cov* (***w***_*t*_) and observation noise covariance ***R***_*t*_ = *cov* (*v*_*t*_) (a scalar in the one-dimensional observation case) we can write the following sequence of steps to perform optimal inference via Kalman filtering mechanism:

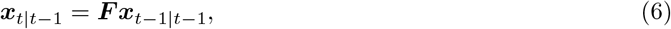

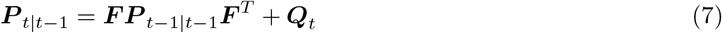

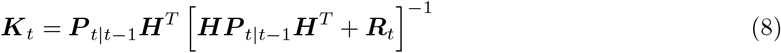

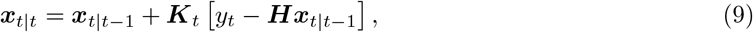

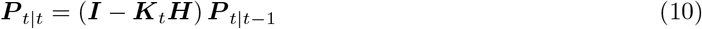

The above steps represent the most general case and allow for optimal inference in the non-stationary noisy environment where driving noise covariance ***Q***_*t*_ and the observation noise covariance ***R***_*t*_ are allowed to change over time. The algorithm then uses Kalman gain ***K***_*t*_ to optimally balance between the *a priory* information about the process conveyed by the state equation and the information present in the current measurement. Often, however, the process and observation noise covariance matrices are assumed fixed. Then Kalman filtering procedure boils down to a very simple recursive estimation scheme termed as steady state Kalman filter (SSKF) :

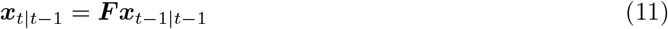

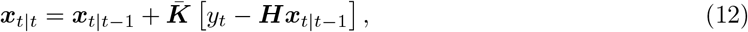

where 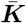 is the steady state Kalman filter gain computed using steady-state state covariance 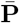 as:

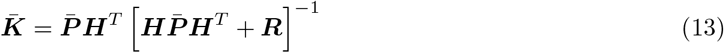

The steady state covariance 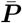 for the eq. 13 can be found by solving once for a set of given parameters the discrete time algebraic Riccati equation:

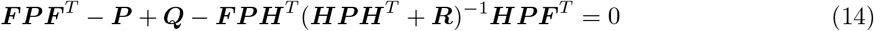

Alternatively, one can obtain 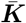 by just letting the filter run for some time before ***K***_*t*_ converges to a steady state value 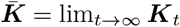.

When operating with typical EEG data pre-processed by a set of hard-coded high-pass and DC-blocking filters running *on-board* of a typical modern EEG machine the *white* KF manages reasonably well and outperforms the existing methods for low-latency phase estimation [15, 53]. At the same time, only this set of hard-coded on-board of a typical EEG device front-end filters typically incurs about 40 ms of delay which in case of beta oscillations amounts for the whole period. We would like to stress here that the filters mentioned above are not the acquisition software filters but what is referred to as “hardware filters” typically implemented in modern EEG devices via digital filtering of the oversampled EEG signal and are often neglected by the researchers. Therefore aiming to provide lowest possible latency feedback based on the parameters of rhythmic EEG components we will minimize these filters to a simple DC-blocker and get confronted with powerful low frequency signal content whose presence adversely affects the performance of *white* KF.

There exist solutions extending KF to operate with colored observation noise *v*_*t*_. Here we have chosen the state-space approach where the noise is modelled as an infinite order AR process driven by a white noise source. In practice a finite but large model order *N* is chosen [28] and the lagged samples of this AR process are then added to the vector of states of our dynamical model which allows us to efficiently account for the 1*/f* nature of noisy components in the raw EEG signal including the contributions of slow potential drifts. See equation 15 describing the *N* -th order AR-process with a set of AR coefficients *ψ*_*i*_, *i* = 0, …, *N* − 1 and *ψ*_0_ = 1.

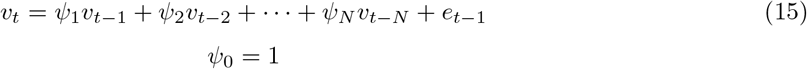

This autoregressive process is known to have the following power spectral density

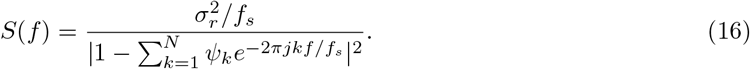

As shown in [28] the auto-regressive coefficients approximating the process with power spectral density (PSD) *P* (*f*) = 1*/f* ^*α*^ can be found with the following recursion

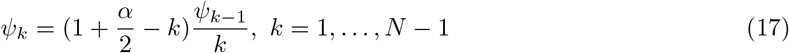

starting with *ψ*_0_ = 1; Parameter *α* is found based on a prerecorded sample of EEG data. Our experience shows that the AR model with order *N* = 100 captures the EEG noise sufficiently accurately starting from 0.5 Hz with sampling rate *f*_*s*_ = 250 Hz, see Figure 1.

**Figure 1.**
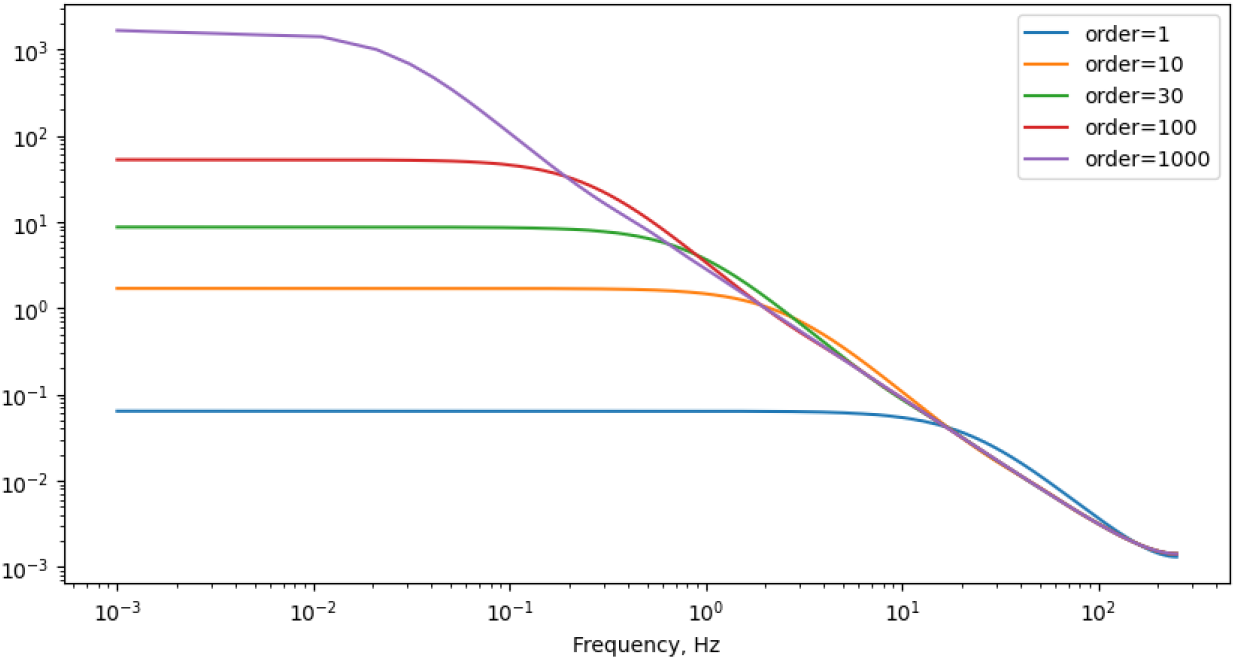
The theoretical spectrum of the truncated 1*/f* ^1.5^ noise autoregressive model for various truncation orders

State space model for the *N* -th order noise will then read as

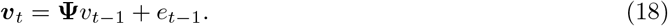

where ***v***_*t*_ = [*v*_*t*_, *v*_*t*−1_, …, *v*_*t*−*N*+1_]^*T*^ is the noise state space vector.

To use this noise state space model in the KF framework we use augmented state space vector 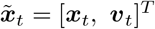 and reformulate the dynamic model 4 and the observation equation 5 as follows

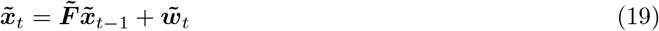

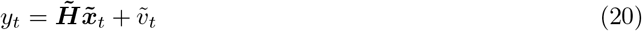

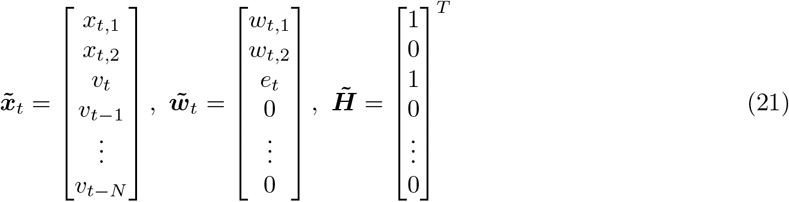

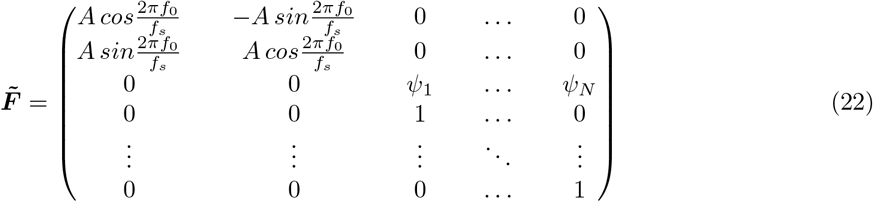

Note that in the limiting case, when *α* = 0 and the AR model order *N* = 1, we get the classical white noise Kalman filter formulation. Importantly, since the observation noise *v*_*t*_ has now become a part of the state model, the noise term 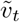 formally present in equation (20) has zero variance. To deal with the associated issues we use the perturbed-P approach shown to be robust and outperforming other forms of colored KF [61].

To facilitate the implementation of this model on board of an EEG device with typically rather modest computational power and available memory resources this state-augmented colored Kalman filter can also be used in the steady state mode. We fit *A, f*_0_, *α, σ*_*q*_, *σ*_*q*_ parameters using the genetic algorithm in a manner similar to that described in [67] and using short segment of prerecorded EEG data from a subject before the main real-time part of the experiment, see also Figure 2.

**Figure 2.**
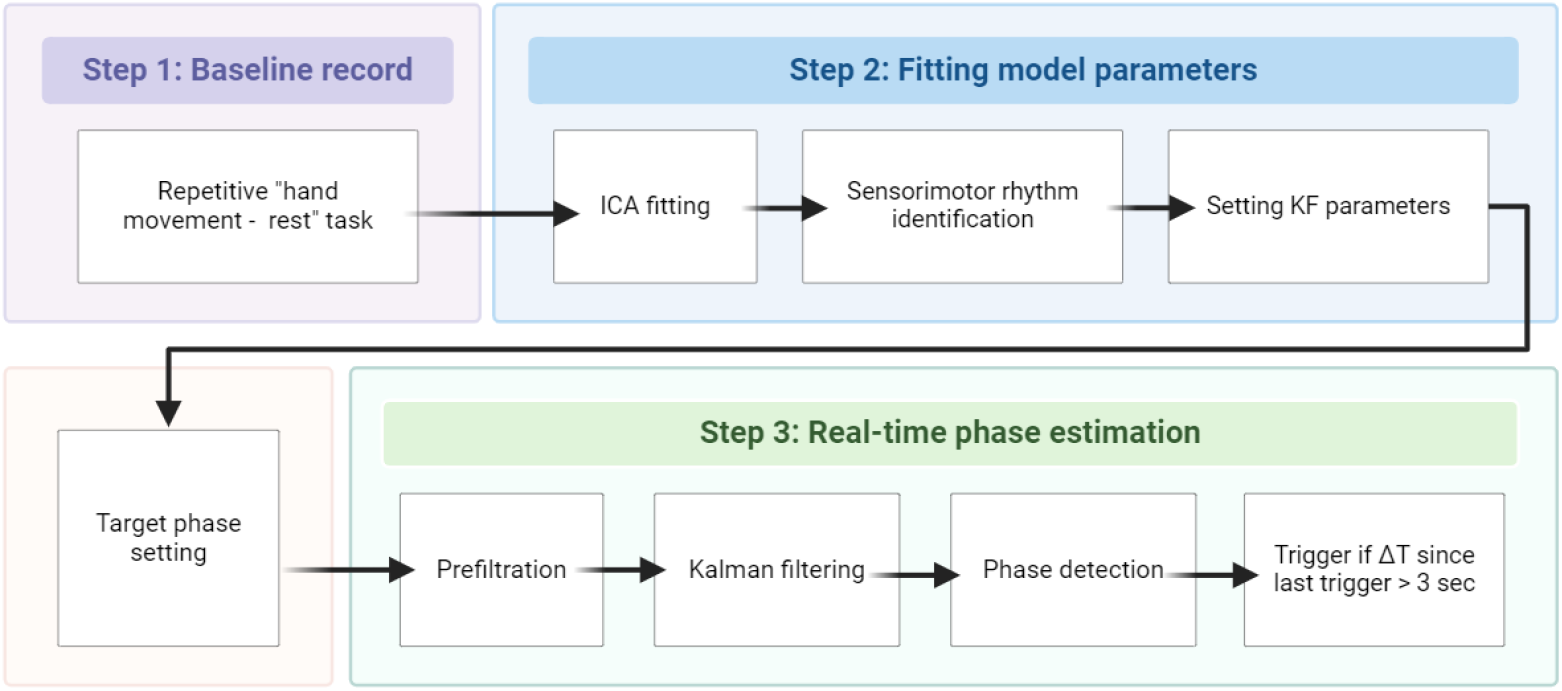
Computational pipeline

### 2.2 Hardware implementation for real-time phase estimation

The main reasons to implement the described algorithms on-board of an EEG device and exploit its truly real-time operation system (OS) are 1) to eliminate the EEG-to-PC data transfer delays [51] and 2) to eliminate variability in computing times due to non-real time nature of an OS controlling a typical PC.

As outlined in Figure 2 our solution comprises two parts. The first part of our application runs on a regular PC and estimates the parameters of our model described in Section 2.1 (Step 2 in Figure 2) using a prerecorded segment of EEG data (Step 1 in Figure 2). The identified parameters are then transferred on-board to the EEG device to enable the SSKF operating on the real-time DSP system perform phase estimation according to (11), (12), (13), (14) as *θ* = Arg (*z*_*t*_) and generate a TTL trigger when the estimated instantaneous phase falls within the user specified ballpark of the target phase values.

The SSKF described in Section 2.1 is implemented on-board of a multichannel scientific and medical-purpose EEG amplifier NVX-52 (Neurovisor) manufactured by Medical Computer Systems LLC (https://mks.ru/). In the core of the NVX-52 is a fixed-point digital signal processor TMS320VC5509A, which allows for high-speed computations, has the DMA-regime, I2C interface, and provides over 300 KB memory. We used integer variables to perform calculations on this 32-bit CPU and for this purpose rescaled the SSKF parameter values and refactored each multiplication and division operation to ensure minimal loss in computational precision. To chose the optimal rescaling and factorization strategy we performed numerous experiments on a regular PC emulating the computations based on integers.

NVX52’s electrodes system allows for a total of 48 EEG channels, but in this work we resorted to using only 5 TMS compatible Ag/AgCl sintered ring electrodes located over the area of sensorimotor cortex: F1, F5, FC3, C1, C5 according to the international 10-10 system, along with a reference electrode aligned with the ground. The electrode impedance values were monitored throughout the experiment and kept below 5*K*Ω. The flat design of the electrodes allows for positioning the coil as close to the head as possible, while the minimum metal content of the electrodes minimizes the artifacts in EEG induced the magnetic pulse.

Nexstim Navigated Brain Stimulation (NBS) System 5 (Nexstim Ltd., Helsinki, Finland, version 5.2.3) was used as a TMS device. The Nexstim NBS system combines non-invasive TMS with MRI-based stereotacsic navigation and simultaneous electromyography (EMG) measurement. It is also equipped with external trigger connectors on the back of the cart. One of the output trigger channels of the EEG amplifier was used to generate a digital output (TTL) pulse to trigger the TMS pulse generation.

It was shown previously [36], that the sensorimotor rhythm has special statistical properties that allow for its reliable extraction using the independent component analysis (ICA). The ICA components are obtained from the EEG data by computing the weighted sum of the EEG channel time series with a set of coefficients stored in the ICA unmixing matrix. Each row of the this matrix stores coefficients to extract a single ICA component time series. To identify the component with a pronounced SMR rhythm we prerecorded a short segment of EEG data during a series of altered intervals of the hand movement and relaxation, (Step 1 in Figure 2). This allowed us to isolate the appropriate ICA component containing the SMR by tracking the expected rhythm’s desynchronization induced by the movement and assessing the component’s power spectral density and spatial topography.

We the used the selected component time series and estimate the parameters of our models along with the steady state KF gain and the associated covariance matrices. The quantities are then passed to the NVX52 amplifier’s DSP. Before the actual experiment starts we also set the target phase value to trigger the TTL pulse. As detailed in Step 3 box of Figure 2 the real-time processing pipeline comprises causal IIR filtering with the 1st-order DC-blocker filter with cutoff frequency of 4 Hz, the 2-order notch 50 Hz filter and the grid of the 1st-order notch filters with stop-bands centered around 100, 150, 200, 250 Hz. These operations are necessary when using white SSKF as its performance is drastically affected by slow potentials with a characteristic time less than 1.0 second and the power line noise 50 Hz. Altogether the pre-filtering leads to the moderate (no more than several milliseconds) group delay due to the mild cutoff frequencies and minimal required orders. The choice of the IIR against FIR filtration is made in order to save on the computing power. Finally, phase estimation is performed according to the state-space model described in Section 2.1. When the estimated phase value falls within the user specified target range, a trigger is generated at the output of the EEG amplifier and passed to the TMS machine. To avoid excessive repetitive stimulation we added a refractory period of 3 s from the last stimulation.

There is also a recent study [39] nicely illustrating an intuitive fact that the oscillation’s phase appears to be a reliable predictor of MEP amplitudes only within the burst of the rhythm. The intuition behind this is the fact that the oscillatory phase is properly defined only when the oscillation itself is present so that the phase is meaningful and can be reliably tracked. HarPULL’s algorithmic core readily supports such envelope contingent triggers as SSKF estimates of the analytic signals furnish direct and algebraically instantaneous access to not only the phase but also to the brain rhythm’s envelop samples. The threshold on the on-line estimated envelope can then be used in combination with the target phase values to condition the trigger generation. At the same time one has to realize that unlike phase estimates whose latency is only about several milliseconds, envelope estimates demonstrate significant lags on the order of several tens of milliseconds [48, 51].

### 2.3 Validation of the HarPULL technology

To verify the validity and reliability of HarPULL’s algorithmic components and its implementation we performed a series of the off-line and on-line tests using both real and simulated data.

#### 2.3.1 Phase estimation accuracy on the simulated data

As a test of HarPULL’s algorithmic stuffing we first assessed the accuracy of phase estimation furnished by the *white* and *pink* KFs and leveraged it against the pioneering and widely accepted PHASTIMATE algorithm used in a range of studies. To this end we simulated the ground truth signal with a linear state-space model described in section 2.1. Additionally we filtered the generated ground truth signal in 8-12 Hz range using the 1-st order Butterworth filter to wean off the spectral tails. We have then added *pink* noise samples with PSD slope parameter *α* = 1.5 which we approximated using the AR model (18) of order *N* = 250. The signal-to-noise ratio (SNR) was set to three different levels: 0.3, 1.0, and 3.0. The examples of simulated signals for different SNR, as well as their power spectral density curves are presented in Figure 3.

**Figure 3.**
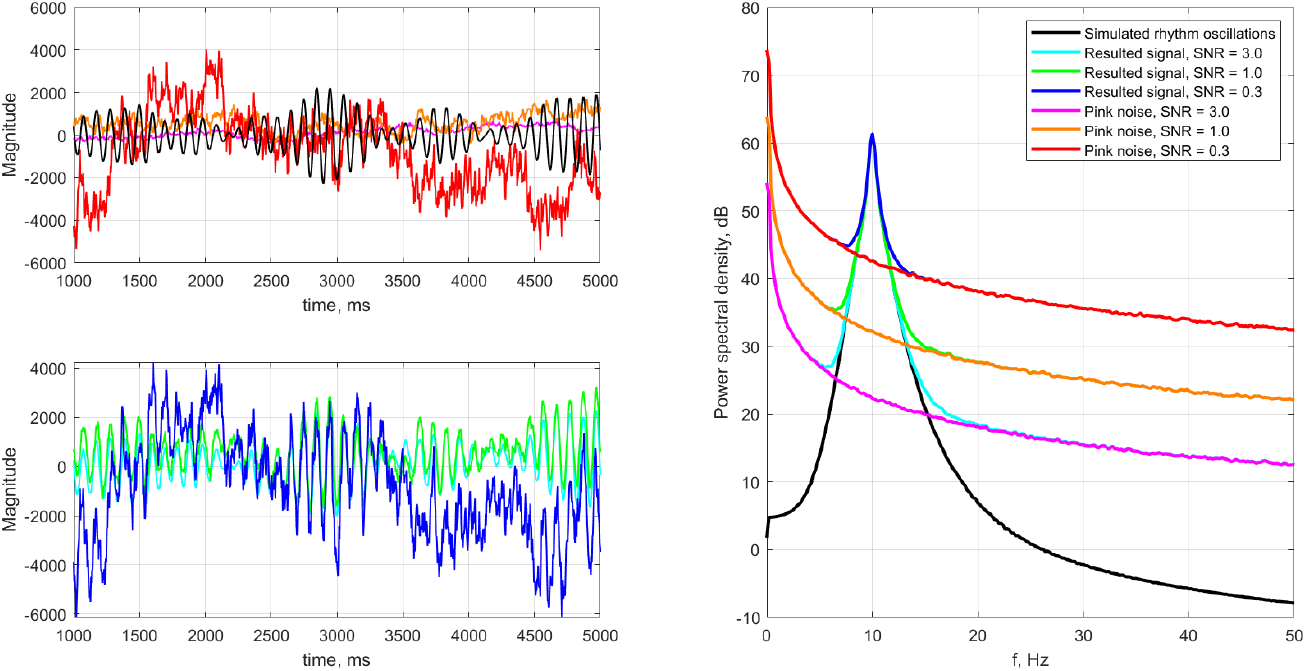
Example 4 s long segments of simulated noiseless time series and noisy observed signals (left) along with their corresponding power spectral density curves (right). See color codes in the right panel

For each SNR value we repeated the simulations 20 times. On each trial using a 4 s long chunk of “training” data we identified the KF parameters as if we were dealing with real data where no ground truth is known. For the *pink* KF optimal parameters were selected by the direct grid search. The parameters of the PHASTIMATE and *white* KF were discovered using the genetic algorithm based approach proposed in [67].

#### 2.3.2 Phase estimation and MEP modulation analysis in real data

We have also applied the three methods (PHASTIMATE, *white* KF and *pink* KF) to a prerecorded real data and assessed the phase estimation error and the depth of phase dependent MEP modulation.

The PHASTIMATE algorithm [67] relies on data from the publicly available dataset accessible at https://gin.g-node.org/bnplab/phastimate and consisting of two parts. In the first part (*murhythmdataset*) the 250 s of the resting-state EEG from total of 140 participants is presented. Within the paradigm adopted by the author, the ground-truth phase is determined at the center of each epoch non-causally using various band-pass filter designs followed by the Hilbert transform. This part of the dataset is used to investigate the phase estimation accuracy.

The second part of the dataset (*mepdata*) contains 1150 one second long segments of scalp EEG recordings from one healthy volunteer synchronized with the TMS-evoked motor responses of the right *abductor pollicis brevis* muscle. All EEG recordings are spatially filtered with a C3-centered Hjorth-style Laplacian and down-sampled to 1 kHz. During the TMS stimulation 160 *µs* long biphasic magnetic pulses were administered using a figure-of-eight coil oriented perpendicularly to the left precentral gyrus with the second phase of the induced electric field in the posterior-anterior direction. The stimulation intensity was 115% of the resting motor threshold [67]. This part of the dataset was used to investigate the level of MEP modulation through the post-hoc sorting of trials, where TMS pulses were applied open-loop at a random phase at the time of the stimulus. As a measure of MEP modulation we used mutual information (MI) assessed by the histogram method between the phase and MEP’s peak-to-peak amplitude. Also similarly to [67] we performed circular regression fit to assess the depth of MEPs modulation.

#### 2.3.3 Real-time phantom experiment

As the next step of HarPULL’s evaluation we performed a real-time test of the *white* Kalman filter using a physical phantom whose electrical activity was driven by a synthetic signal. The *white* KF was chosen for the real-time application due to the combination of its simplicity, super modest computational requirements and a very decent performance in the phase-tracking task, see [63] and our comparisons reported further in Section 3.

A medium-sized ripe watermelon was taken as a phantom (Figure 4). We used open-source electronics platform Arduino Mega 2560 and its DAC as a signal generator. We modelled “brain’s” rhythmic activity according to the linear state-space model equation 4 and stored it as the ground truth for further calculations. To input this signal into the watermelon we used needle contacts inserted into watermelon’s rind. HarPULL modified EEG amplifier NVX-52 was used for recording the electrical fields of the watermelon induced by the generator. To this end, two EEG electrodes (recording and reference) were placed on the pre-treated with alcohol wipe section of the rind so that the impedance under each electrode was maintained at less than 5*k*Ω.

**Figure 4.**
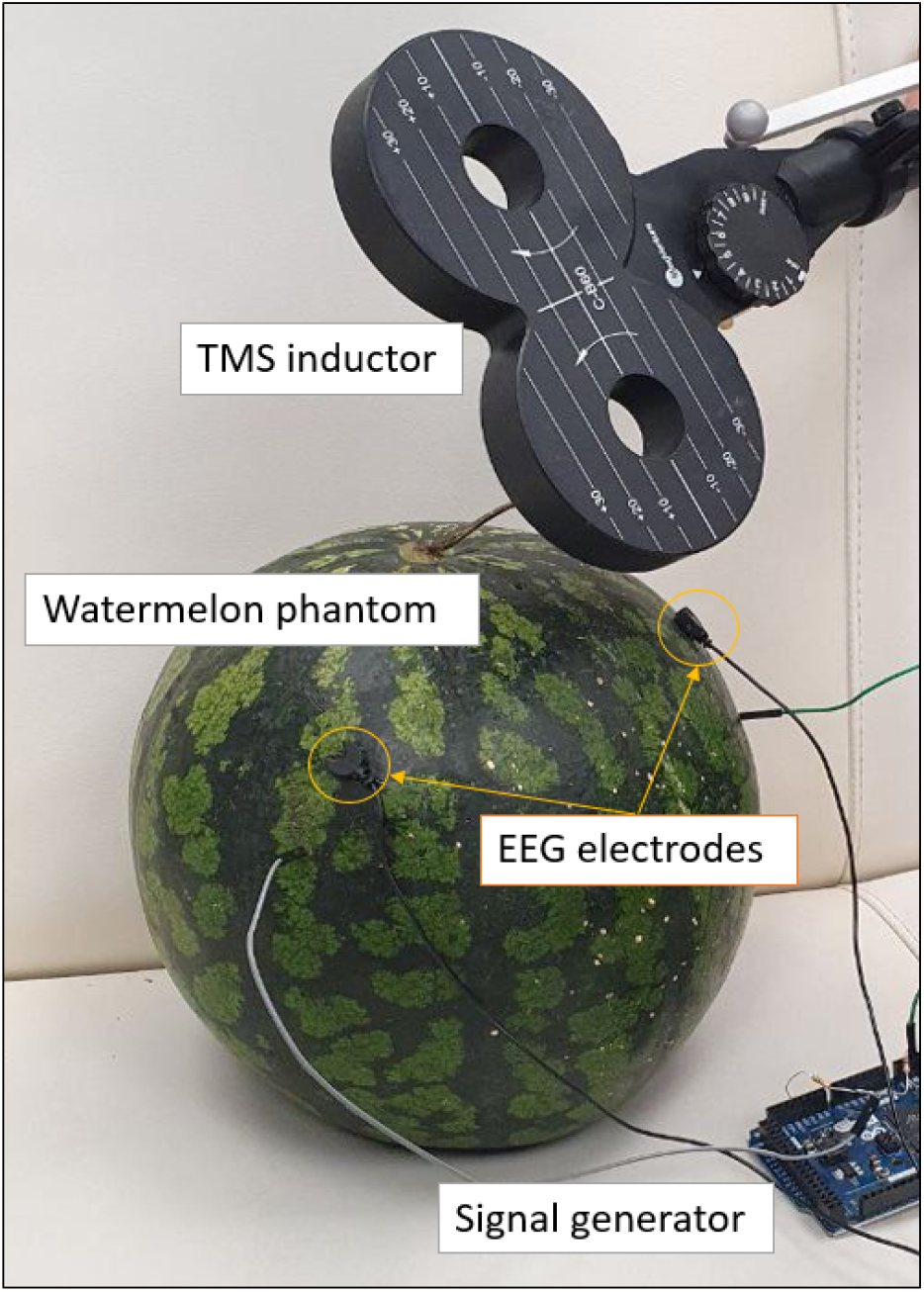
A watermelon phantom model setup

Again, we prerecorded a short segment of data in order to estimate the parameters of the Kalman filter to transfer this information to our phase tracking algorithm running on board of NVX-52 EEG amplifier. Spectral analysis was performed to determine the peak frequency and the SNR of the modelled resultant signal which was necessary due to the presence of additional technical noises.

The TMS system was triggered by the HarPULL NVX-52 EEG amplifier when the extracted signal phase appeared in the ball-park of the pre-determined target phase value, see Figure 2. To sample the entire range of phase values we have randomly chosen the target phase value from the [− 180°,180°] range split into 20° intervals. Trigger formation moments were recorded in a separate EEG channel and stored at Arduino Mega 2560. The moments of magnetic stimulus arrival on the rind were recorded by the Arduino’s board registration channel with zero latency. At the moment of trigger formation the inputs of the amplifier were switched off to avoid excessive saturation due to the following TMS pulse. Thus, on Arduino’s side we have the ground truth signal, the target phases and the moments of time, when triggers arrived to the watermelon rind. This allowed us to calculate the overall delay from the moment of the desired event (“Arduino GT signal”) in the watermelon rind (target phase) and the moment of trigger arrival to the registering channel in the rind and input back into the Arduino device (“Detected Phase”). The corresponding delay measurement diagram is shown in Figure 5.

**Figure 5.**
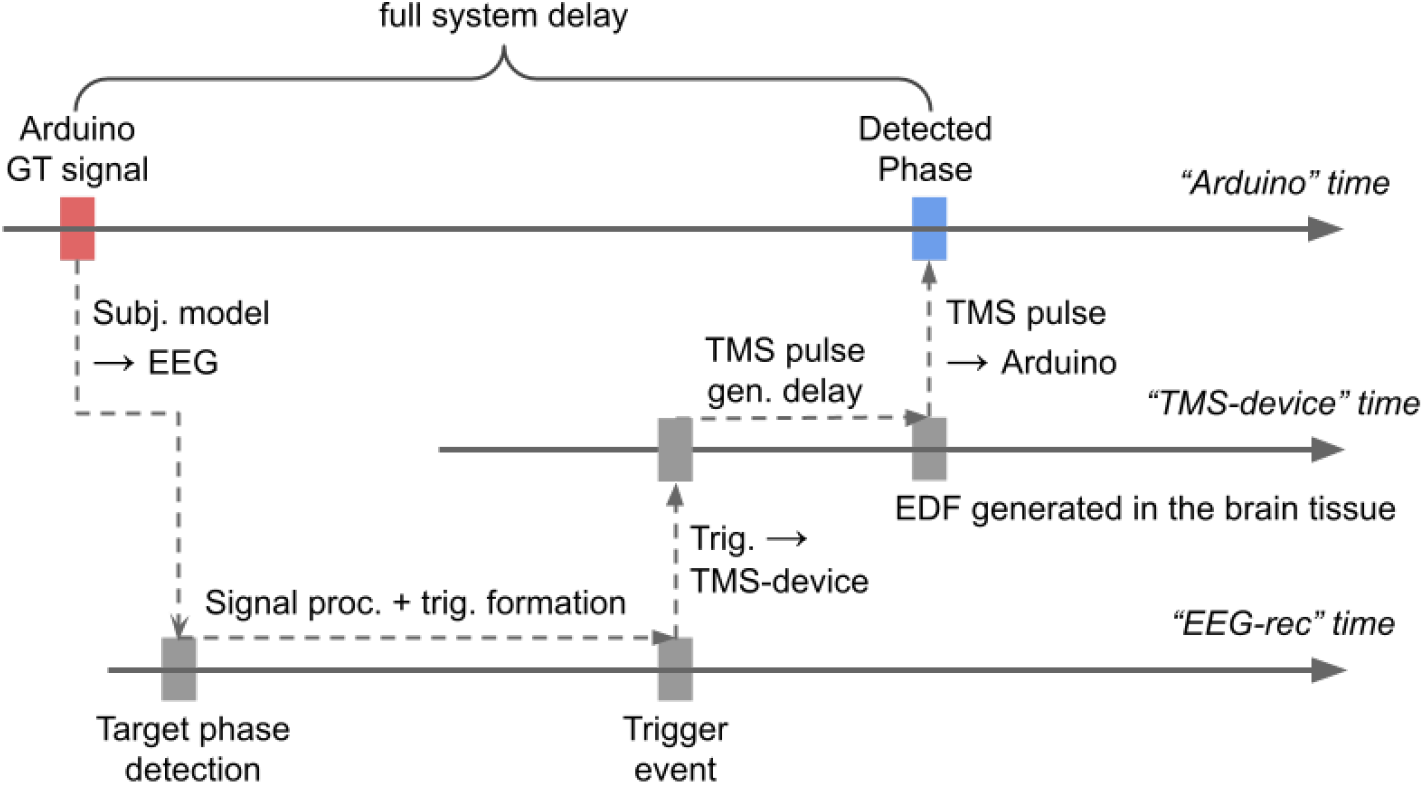
Latency measurement diagram. We used an Arduino’s DAC to generate the analog rhythmic signal simulated in agreement with the state space model 4. This signal was then applied to watermelon and input to HarPULL’s EEG device where the phase tracking algorithm was run to detect the target phase moment and generate a stimulus to be sent to the TMS stimulator. The TMS simulator in response to the trigger signal generates a magnetic pulse that is also applied to the watermelon and induces a splash of electrical activity registered back by the Arduino’s ADC. The latency is measured as the delay between the target phase moment (“Arduino GT signal” time stamp) and the moment when the TMS device generated a TMS pulse, see “Detected Phase” time stamp on the Arduino’s timeline.

#### 2.3.4 Real-time healthy volunteer experiment

To verify the real-time operation of the system we conducted an experiment according to the protocol shown in Figure 6. The protocol has been prepared and implemented to agree with the earlier published guidelines for state-dependent TMS [20, 54].

**Figure 6.**
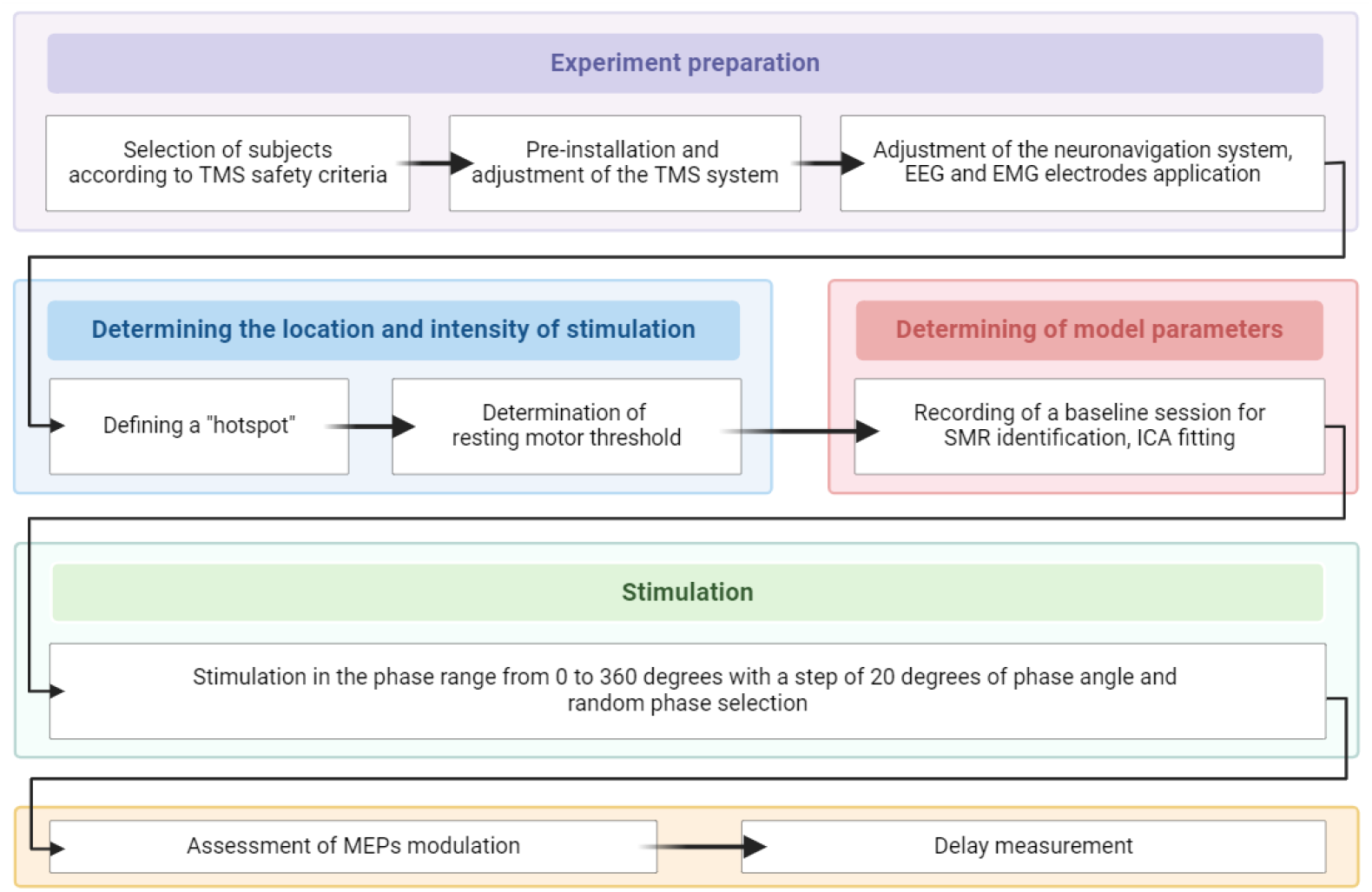
Real-time healthy volunteer experimental protocol

A male subject with no neurological and/or psychiatric impairment and no contraindications to TMS according to the screening questionnaire (modified by Rossi et al, 2009) [42], who abstained from alcohol consumption at least two days before the experiment and nicotine and caffeine for at least four hours before the experiment, with a pre-recorded structural MRI of the brain was seated in a comfortable position, avoiding head and arm movements as much as possible. The subject was instructed to avoid the behaviors inducing typical EEG artifacts (e.g. chewing, jaw clenching, yawning, or talking) throughout the experiment.

Further preparation included checking the equipment operability, setting up the navigation system, placing EMG electrodes according to the “belly-tendon” rule on the selected muscle, in this case m. abductor pollicis brevis (APB) and EEG electrodes. The early electrode placement was aimed at setting the stimulation parameters considering the “peeling” determined by the presence of the EEG cap. The optimal “hotspot” locus was found with 80-points rough mapping, the corresponding coil position (its orientation and angle) was stored in the memory of the neuronavigation system. Resting motor threshold (RMT) was then defined using an automated protocol built into the NBS software [42]. The TMS was performed with a power of 110% of the found RMT. The phases of SMR at which the stimulus was applied, were randomly selected from a series formed by values from 0 to 360 in steps of 20 degrees. A total of a 100 pulses were delivered that is sufficient to manifest the effect of MEP modulation [67].

#### 2.3.5 Navigated TMS (nTMS) motor mapping

Muscle cortical representations (MCRs) of different muscles can also be assessed using state-dependent TMS approaches. Changes in the amplitude of motor responses depending on the phase of the sensorimotor rhythm are hypothesized to lead to changes in key mapping metrics such as center of gravity (CoG) (the location on the grid most likely to produce the largest MEP), area (the spatial extent of a motor map in *cm*^2^) and volume (the MEP amplitude weighted spatial extent of a motor map in *mV cm*^2^) [52]. To explore the effects of the real-time phase dependent nTMS mapping, we constructed MCRs for the four muscles: abductor pollicis brevis (APB), abductor digiti minimi (ADM), extensor digitorum communis (EDC), and biceps brachii (BB) at rest in another male right-handed healthy volunteer.

The subject did not have specific fine motor skills such as playing musical instruments, hobbies, or occupations that constantly involve them. He also met the criteria and had no contraindications to stimulation according to the modified screening questionnaire [42]. Prior to participation in the experiment, the subject underwent an MRI procedure to obtain structural scans required for a neuronavigated TMS. Five recording electrodes (C3, CP1, CP5, FC1, and FC5) were placed on the subject’s head according to the international 10-10% system, and impedance was maintained at *<* 5*kOhm*. We verified the absence of activation by surface EMG prior to delivery of the TMS pulse. The hotspot and RMT were determined similarly to the previous experiment (see Section 2.3.4).

We implemented the mapping approach using the grid with 5×5 mm spacing centered on the hotspot defined earlier. The mapping was performed with single pulses of 110% of the defined RMT. The points on the grid were placed as far apart as possible in a randomized order to avoid prolonged stimulation of the same area. A mapping session was performed until a closed contour was formed around the map by points with MEP amplitudes less than 20 uV.

We repeated the mapping for three different conditions: in the state of maximum excitation of the neuronal ensemble (target phase value 180°), which corresponds to the trough of the sensorimotor rhythm, in the state of maximum inhibition (target phase value 0° corresponding to the sensorimotor rhythm’s peak) and in the random phase of the sensorimotor rhythm. In addition to the obtained maps created with *TMSmap* software [38] (https://tmsmap.ru/) we calculated the MCR area as a measure of change in motor representation maps.

#### 2.3.6 Phase tracking error definition

To ease interpretation we used the circular standard deviation of the difference between the estimated and true phase values, as proposed by Zrenner et al. [67]:

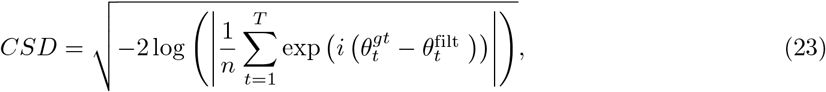

where 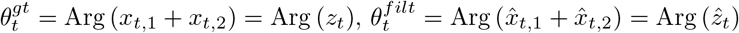 are ground truth phase and phase of the estimated state at the time moment *t* respectively; *n* is a number of samples.

## 3 Results

We first examine the accuracy of phase estimation with *white* and *pink* KF approaches on the simulated and real data and compare it against the most common method proposed by Zrenner et al. (PHASTIMATE) [67]. We then illustrate real-time HarPULL’s performance withing the state dependent TMS experiments with a phantom and a healthy volunteer. In the phantom experiment where we know the ground truth we explore the overall latency of our system, i.e. the time that lapses between the “neuronal” event (Arduino simulated rhythm’s passing through specific phase) and the TMS pulse. In the first experiment with a healthy volunteer we examine the modulation of MEP amplitude by the target SMR’s phase value. In the second experiment with a healthy volunteer we build the MCRs for four muscles and show how the phase dependency affects these MCRs.

### 3.1 Phase-estimation error on simulated data depends on the SNR and minimal when using *pink* Kalman filter

We used the two KFs and PHASTIMATE approaches to track phase in the data simulated according to the strategy described in Section 2.3.1.

In Figure 7.a for each SNR value we show phase error histograms superimposed for the three methods. Judging based on the mean value of the phase estimation error, see also Table 1, we can conclude that all three methods operate at zero-latency or close to it. PHASTIMATE is characterized by the largest phase delay for all three SNR values which nevertheless even in the worst case scenario at SNR = 0.3 remains below 9°. In the algorithm proposed by Zrenner et al. [67] this is achieved by the use of the auto-regressive model to forecast the narrow-band filtered signal for 150-300 ms to compensate for the filter’s group phase delay. The state-space modelling approach utilised by the Kalman filters deals with the true signal and bases its phase prediction on the physiologically plausible model of the brain rhythm as a frequency modulated process proposed by N. Wiener [62]. Also, both Kalman filters require fewer parameters to be estimated from the data than PHASTIMATE. Figure 7.b shows a graphical summary of the phase tracking error analysis in the form of the phase estimation error CSD, see equation (23) for the three methods and three SNR conditions. As we can see, *pink* Kalman approach noticeably outperforms the *white* Kalman filter and the PHASTIMATE technique. The performance improvement furnished by the more complicated state-space model of the *pink* Kalman is most pronounced in the lowest and the highest SNR cases. Modelling actual noise dynamics afforded by Kalman filtering yields fastest improvement of phase tracking performance with the growth of the SNR. Note that the presented results were obtained on the basis of the noise and rhythm model parameters estimated from the corresponding data sample (see section 2.3.1, similarly to the way it was done when dealing with real data and not using the true set of signal and noise parameters typically not available in real life.

**Table 1.** Mean errors and CSD values for PHASTIMATE, *white* and pink KF with different values of SNR.

| Method | Mean error/CSD |  |  |
| --- | --- | --- | --- |
|  | SNR = 0.3 | SNR = 1.0 | SNR = 3.0 |
| PHASTIMATE | 8.36°/55.17° | 4.63°/39.46° | 0.95°/36.67° |
| *White* KF | 2.4°/48.65° | -0.14°/34.0° | 0.14°/23.78° |
| *Pink* KF | -0.11°/31.70° | -1.53°/20.3° | -0.07°/11.95° |

**Figure 7.**
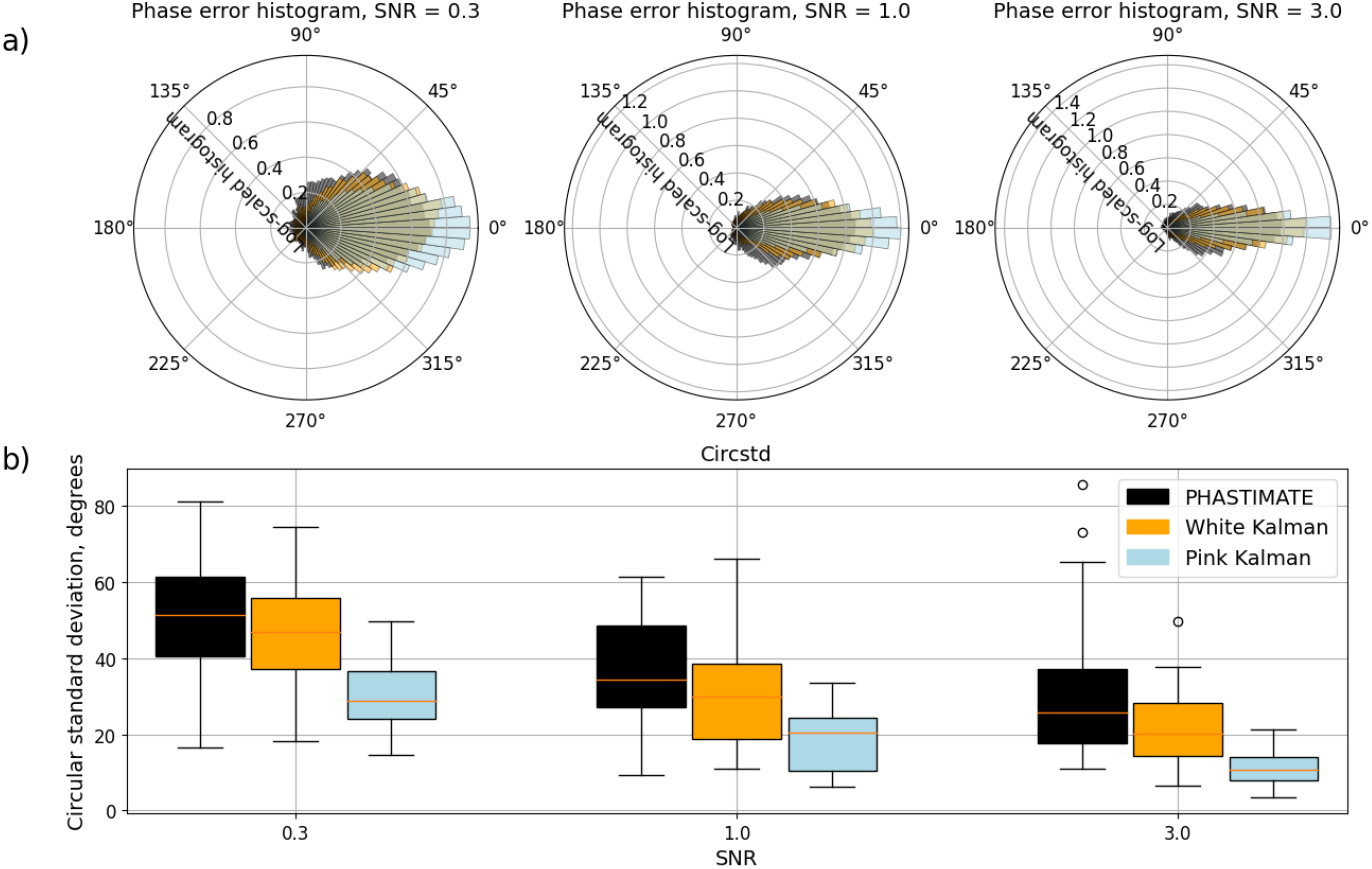
Phase tracking error for the three methods and three SNR values: a) logarithm of the CSD probability density function, b) CSD boxplots

### 3.2 Pink Kalman filter outperforms PHASTIMATE in phase-tracking on real data from murhythmdataset

We applied the three methods to the *murhythmdataset* presented in [67]. Figure 8.a visualizes the overall error distribution including all trials in all subjects for PHASTIMATE, *white* and *pink* KF. The mean error (equivalent to delay) for PHASTIMATE constitutes about 2.8°, for *white* KF – 6.5°, and for *pink* KF - 3.2°. Mean circular standard deviation computed according to equation (16) on the entire dataset is *CSD* = 46.8° for PHASTIMATE, *CSD* = 43.5° for *white* KF, and *CSD* = 43.0° for *pink* KF. None zero mean phase estimation error does not cause problems when operating in real-time as the target phase values can be adjusted accordingly. More important are the properties of the CSD distribution as they reflect the algorithm’s accuracy of hitting the target phase in a real-time setting. In Figure 8.b. we plotted the distribution of the CSD values computed using the bootstrap sampling from the existing trials of the *murhythmdataset*. Visual analysis shows that the lowest mean CSD pertains to the *pink* KF and it appears significantly smaller as compared to the mean CSD of the PHASTIMATE method (ANOVA, *p <* 0.02). Modelling colored noise by the *pink* Kalman filter visually improves performance as compared to the *white* Kalman filter but the difference does not appear to be statistically significant. Overall, the state-space modelling approach exercised by the Kalman filter when the care is taken to accurately model not only the target signal but also the additive noise makes real-time phase estimators align better with the instantaneous phase values obtained using non-causal filtering and not available in real-timee. At the same time, the difference between the performance of the three methods is not as pronounced as it is in the case with real data. Possibly, the “shaky” nature of the ground truth, see [67], leveled out the clear advantages of the state-space modelling approaches observed in the simulated data, see Figure 7, where the ground truth was known.

**Figure 8.**
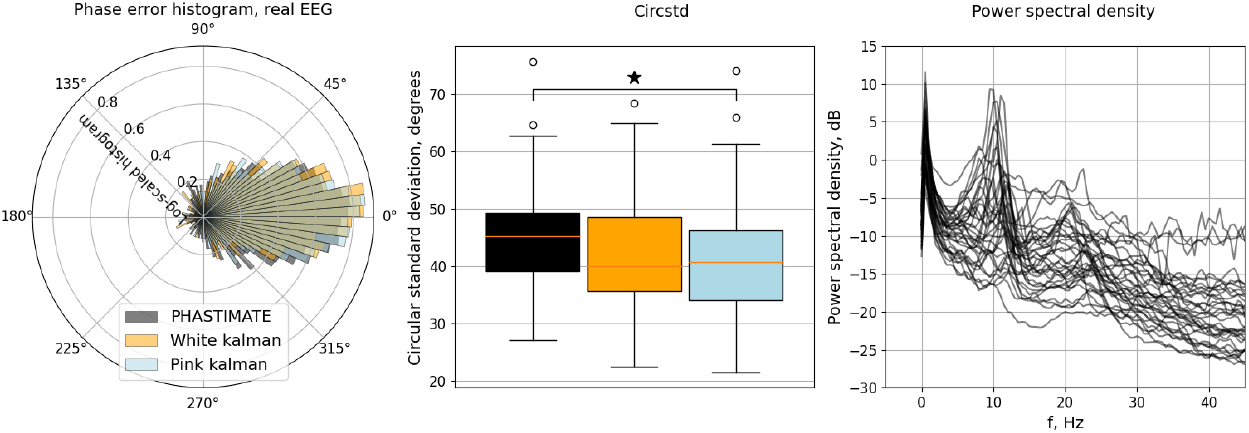
Phase estimation error observed in real subject data from [67]: a) log-scaled histogram of the phase estimation error for the three methods, b) box plot of the CSD for the three methods, c) power spectral density for the data segments supplied by [67]

### 3.3 Sensorimotor rhythm’s phase extracted by all methods significantly modulates MEP amplitudes

The post-hoc analysis of MEP amplitudes as a function of the estimated phase corresponding to the stimulation moment performed on *mepdata* [67] showed that MEP size is larger when the TMS is applied at the negative peak (180° phase) of the sensorimotor rhythm, and smaller when the TMS pulse is aligned with the positive peak (0° phase).

Figure 9 illustrates the MEP amplitudes in log scale vs. estimated phase at the moment of stimulation. Circular-to-linear regression analysis between the estimated phase and the log-transformed MEP amplitudes showed a highly significant correlation with the sinusoidal model with *p* = 10^−32^ for PHASTIMATE, with *p* = 10^−34^ for *white* and *p* = 10^−27^ for *pink* KF.

**Figure 9.**
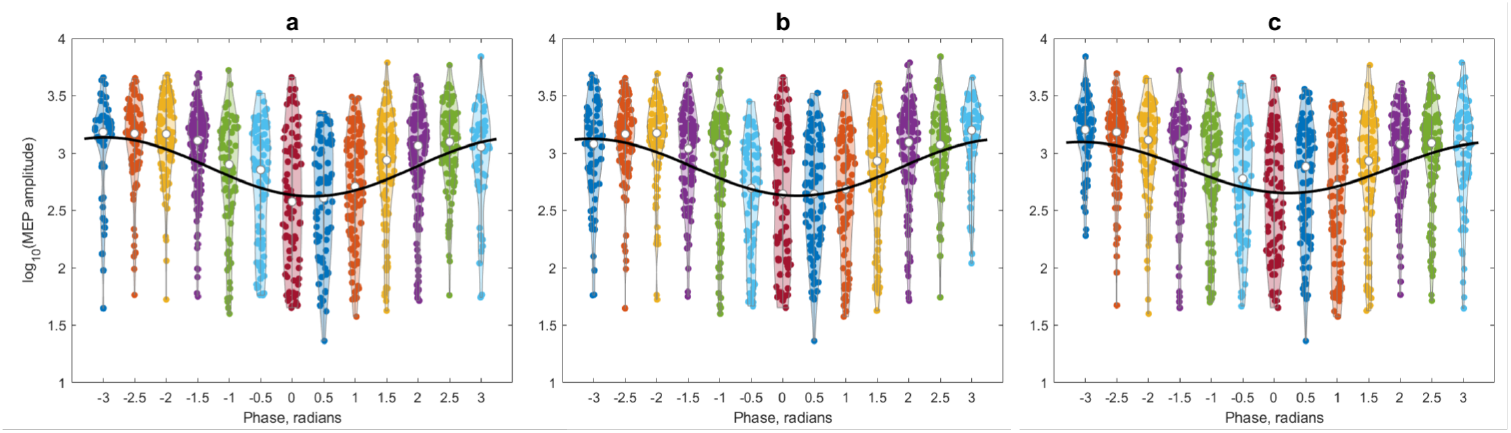
MEP amplitudes (in log scale) and the result of the circular regression fit (black curve) as a function of the stimulation moment phase estimated: a) with PHASTIMATE, b) *white* KF and c) *pink* KF.

To quantify the amount of statistical connection between the MEP amplitude and sensorimotor rhythm’s phase we computed normalized mutual information value between these two quantities. We used histogram method and adjusted bin size to accommodate on average 5 measurement points per bin. All three methods yielded nearly identical value of the normalized MI: *MI*_*n*_ = 0.582 for PHASTIMATE and *MI*_*n*_ = 0.586, *MI*_*n*_ = 0.587 for *white* and *pink* KF respectively. We have also used this MI criterion to optimize parameters of all three methods.

Interestingly, the improved phase tracking performance of the *pink* KF observed with simulated and partly real data does not translate into the improved MEP modulation depth. This may potentially be due to the atypically high SNR of the sensorimotor rhythm in the data at hands [67].

### 3.4 Real-time phase estimation is accurate and stimulus delay is ultra low in a real-time phantom experiment

For a real-time phantom experiment we assembled the hardware and software system as described in Section 2.3.3. The tested system implements steady-state *white* Kalman Filter as a method for tracking the instantaneous phase. NVX-52 hardware and the on-board processor impose significant constraints on the computational complexity of the phase tracking algorithms. Considering this and based on our own experiments comparing the performance of the three phase tracking methods reported above we have chosen *white* Kalman filter to be implemented on board of HarPULL’s NVX-52 EEG device. More advanced signal processing architectures are likely to afford not only the *pink* Kalman Filter but also more demanding neural network based algorithms, e.g. [48] specifically designed for tracking phase and amplitude of brain’s rhythmic activity.

The delay from the moment when the target phase is reached to the moment of the TMS pulse arrival at the recording channel can be calculated according to the phase-detection error histogram, presented in Figure 10.a. Thus the overall phase delay between the TMS stimulation artifact registered on the watermelon’s rind and Arduino’s target ground truth phase value is found to be -2.17 ms. The negative sign of the delay is explained by the effect of the high-pass filter with high cut-off frequency (7 Hz, see section 2.2).

**Figure 10.**
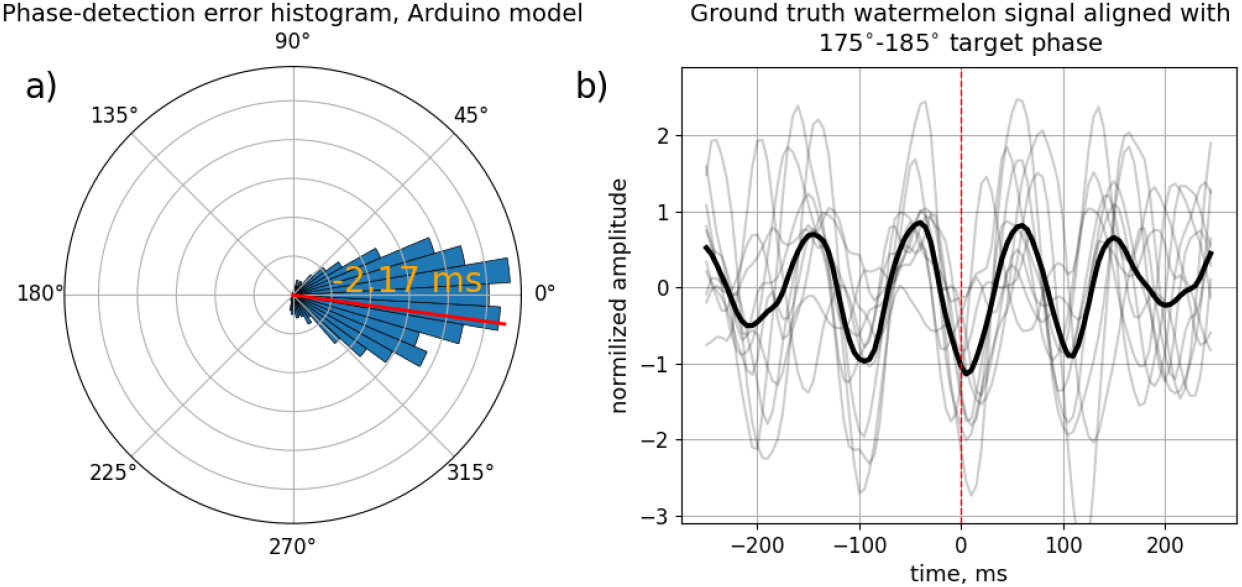
Phase and delay estimation in phantom model: a) phase-error histogram. Red line indicates mean phase detection error - overall latency of the phase-detection system. b) epochs of the signal with target phase 180° aligned with the time ground truth phase moment.

The SNR, determined according to [67] was found to be 1.2. Figure 10.b demonstrates epochs of the ground truth signal aligned at the target phase 180°, which corresponds to the trough of the sensorimotor rhythm. Mean error in real-time phantom experiment constitutes − 7.8° and mean CSD = 31.4°. This observed in the real-time experiment mean CSD value for the estimated SNR = 1.2 matches well the purely computational simulation results reported in Figure 7.

### 3.5 The HarPULL hardware-software complex can be successfully used in the brain-state dependent TMS paradigm

Figure 11.a presents the log-scaled MEP amplitudes (blue dots) and the result of the circular regression fit (red sinusoidal curve), obtained during real-time experiment on a healthy volunteer with 100 TMS pulses applied according to the process listed in Section 2.3.4. A slight decrease in the level of MEP modulation compared to *mepdata* offline experiment can be noted.

**Figure 11.**
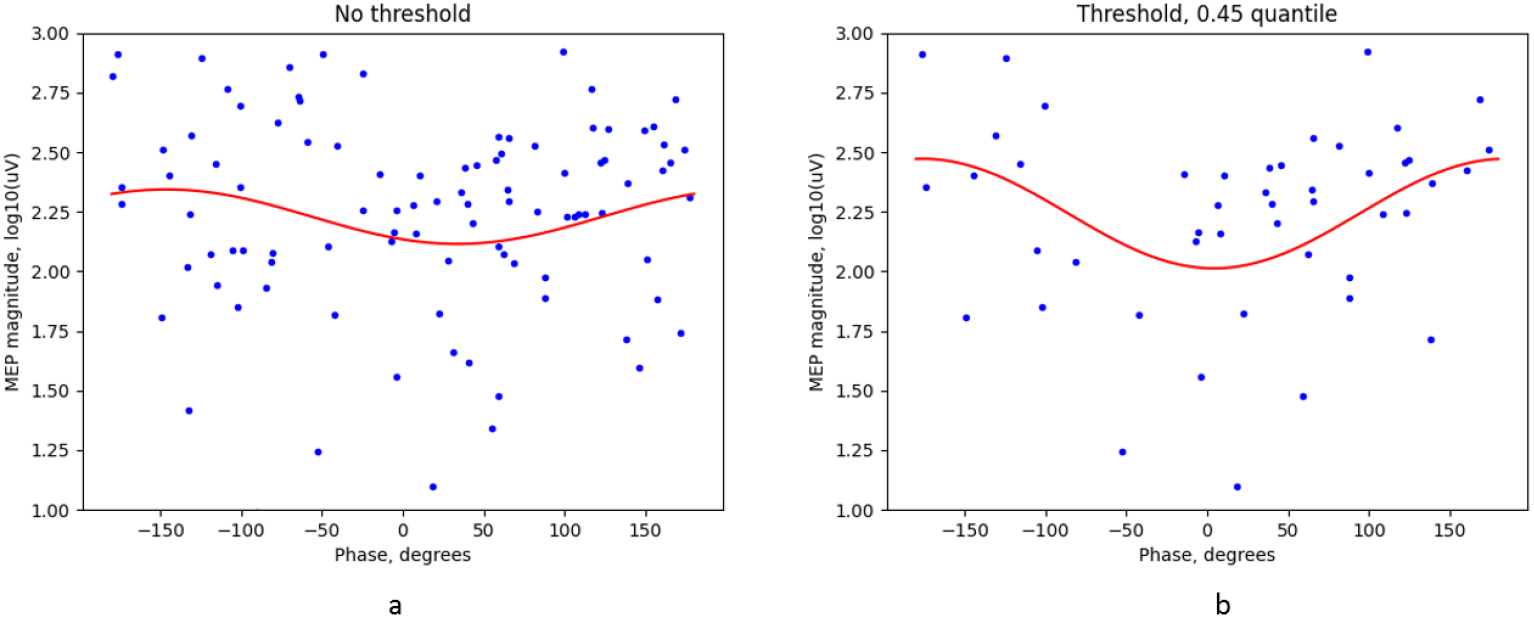
Results of the real-time TMS experiment: a) log-scaled MEP amplitudes (blue dots) and the result of the circular regression fit (red curve) a) without the SNR based amplitude threshold and b) using the threshold corresponding to the 45% SNR amplitude quantile.

It was demonstrated [39] that the error of phase estimation is strongly affected by the rhythm’s instantaneous amplitude. We confirm this in the post-hoc analysis of the data obtained in the real-time experiment by introducing the SNR threshold corresponding to the 45% quantile of the SMR’s burst amplitude threshold. Figure 11.b shows the improved depth of MEP amplitude modulation corresponding to the 55% of most powerful bursts of the sensorimotor rhythm. Within the state-space approach exercised by the Kalman filter our states are the real and imaginary parts of the analytic signals corresponding to the brain’s rhythm of interest, see equation 4. This gives us instantaneous access to the amplitude and therefore the thresholding can be implemented in real-time which further improves the specificity of the state dependent TMS by linking it more tightly to the brain’s functional system state conveyed by its oscillatory activity.

### 3.6 Muscle specific cortical representations are modulated by the phase of sensorimotor rhythm

Finally, we used our real-time HarPULL setup to obtain muscle cortical representations (MCRs) and their areas [5] for four muscles following the methodology described in Section 2.3.5. The results are presented in Figure 12 and Table 2. To the best of our knowledge it is for the first time that the phase dependent MCRs are obtained within a real-time experiment. The demonstrated phase dependency was achieved with much fewer TMS pulses than it would be possible with the traditional approach using post-hoc analysis of the EEG + MEP data recorded in the phase agnostic mode.

**Table 2.** MCR areas and maximum MEP amplitudes for the four upper limb muscles.

| SMR phase | MCR areas* |  |  |
| --- | --- | --- | --- |
|  | phase-neutral | 0°(exc.) | 180°(inh.) |
| APB | 0.95 | 1.25 | 0.95 |
| ADM | 0.92 | 1.45 | 1.01 |
| EDC | 1.01 | 2.13 | 1.33 |
| BB | 0.86 | 1.46 | 0.75 |
| SMR phase | Maximum MEPs** |  |  |
|  | phase-neutral | 0°(exc.) | 180°(inh.) |
| APB | 2662.4 | 3301.2 | 2315.1 |
| ADM | 1129.5 | 682.4 | 1255.6 |
| EDC | 3497.2 | 3419.9 | 1586.0 |
| BB | 656.3 | 458.7 | 788.6 |

*Note.* Phase-neutral - classical nTMS mapping approach without taking into account the SMR phase, 180° - the excitation and 0° - the inhibition states of the motor system.
\* MCR area in $cm^2$ (Gaussian), \*\* maximum MEP amplitudes in $\mu V$

**Figure 12.**
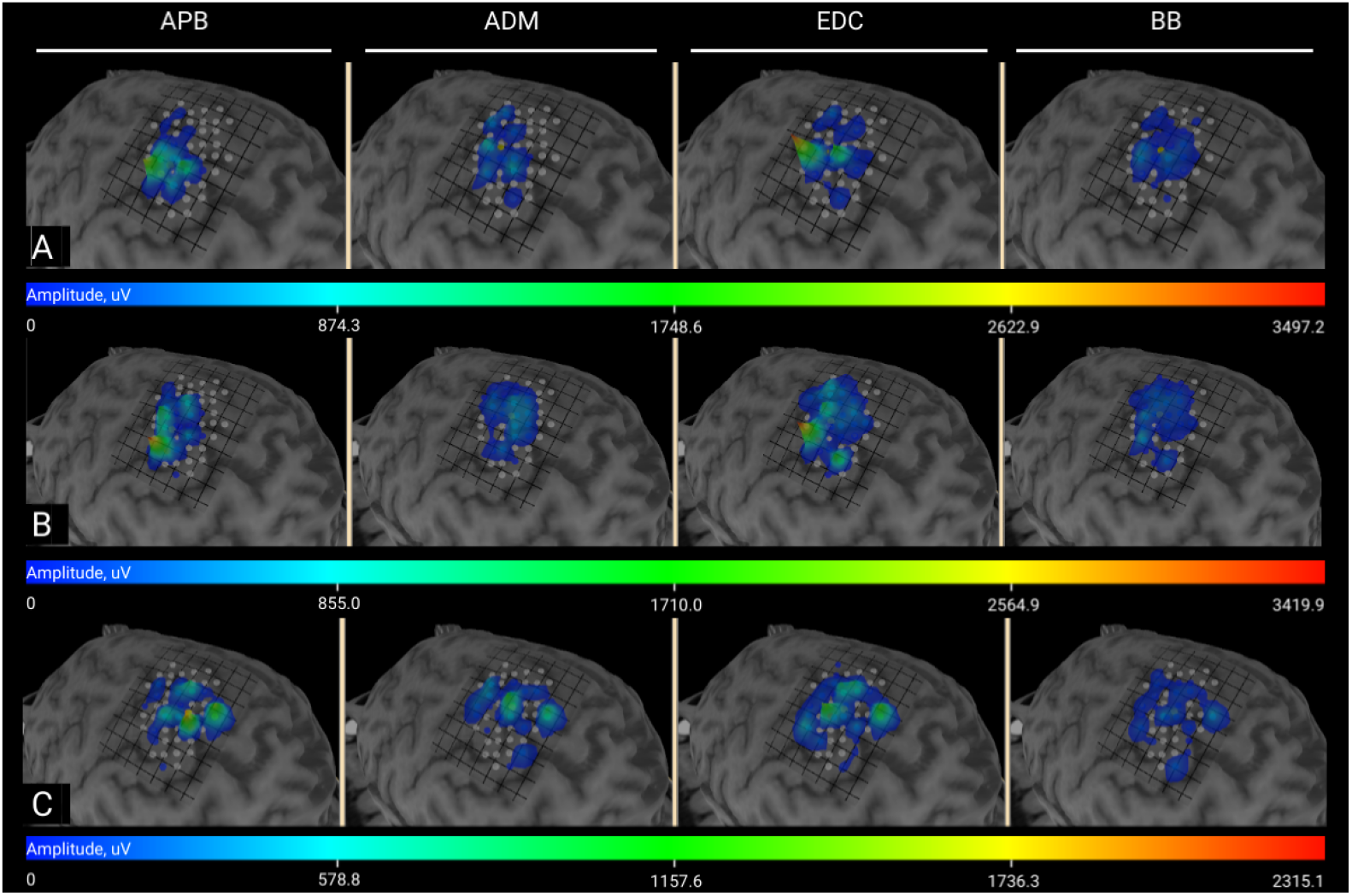
MCR maps: A - phase-neutral stimulation, B - stimulation during the excitation state (*ϕ*_*SMR*_ = 180°), C - stimulation during the inhibitory state (*ϕ*_*SMR*_ = 0°) phase of sensorimotor rhythm

The first row of images, Figure12.a, illustrates results of the traditional TMS stimulation agnostic with respect to the ongoing SMR’s phase. The second and the third rows represent similar maps but obtained in real-time when the TMS pulse was locked to the excitation (panel b, *ϕ* = 180°) and the inhibition (panel c, *ϕ* = 0°) states of the motor system. The color encodes MEP amplitudes according to the color-bars supplied in the bottom of each MCR panel. Note that the scales differ between panels a), b) and c).

First of all, we can clearly see that the MEP for all muscles on average are significantly lower in the inhibitory state (panel c) as compared to the phase agnostic (a) and the excitation (b) states. Interestingly, in the excitation state (b) cortical representation of *Abductor pollicis brevis* appears clearly delineated from that of *Abductor digiti minimi* while such a split is not observed during either phase agnostic (a) or inhibitory mode (c) mappings.

Overall, visual analysis of MCRs shows the largest diversity of spatial maps in the excitatory (b) state as compared to the two other conditions. As shown in Figure 13, the area of the resulting maps varies depending on the phase of the rhythm at which the mapping is performed and is largest at 180° phase corresponding to the excitation state. At the same time, maximum MEP values presented in Table 2 appear to be not mechanistically consistent with the the excitation/inhibition dichotomy. One of the reasons for this could be the fact that EEG based sensorimotor rhythm signal is formed by large neuronal populations which leads to the expected of the integral indicators such MCR’s area. This global nature of the EEG based observations results in its failure to capture the peculiarities of the motor cortex and track the excitation/inhibition of the compact MCR components.

**Figure 13.**
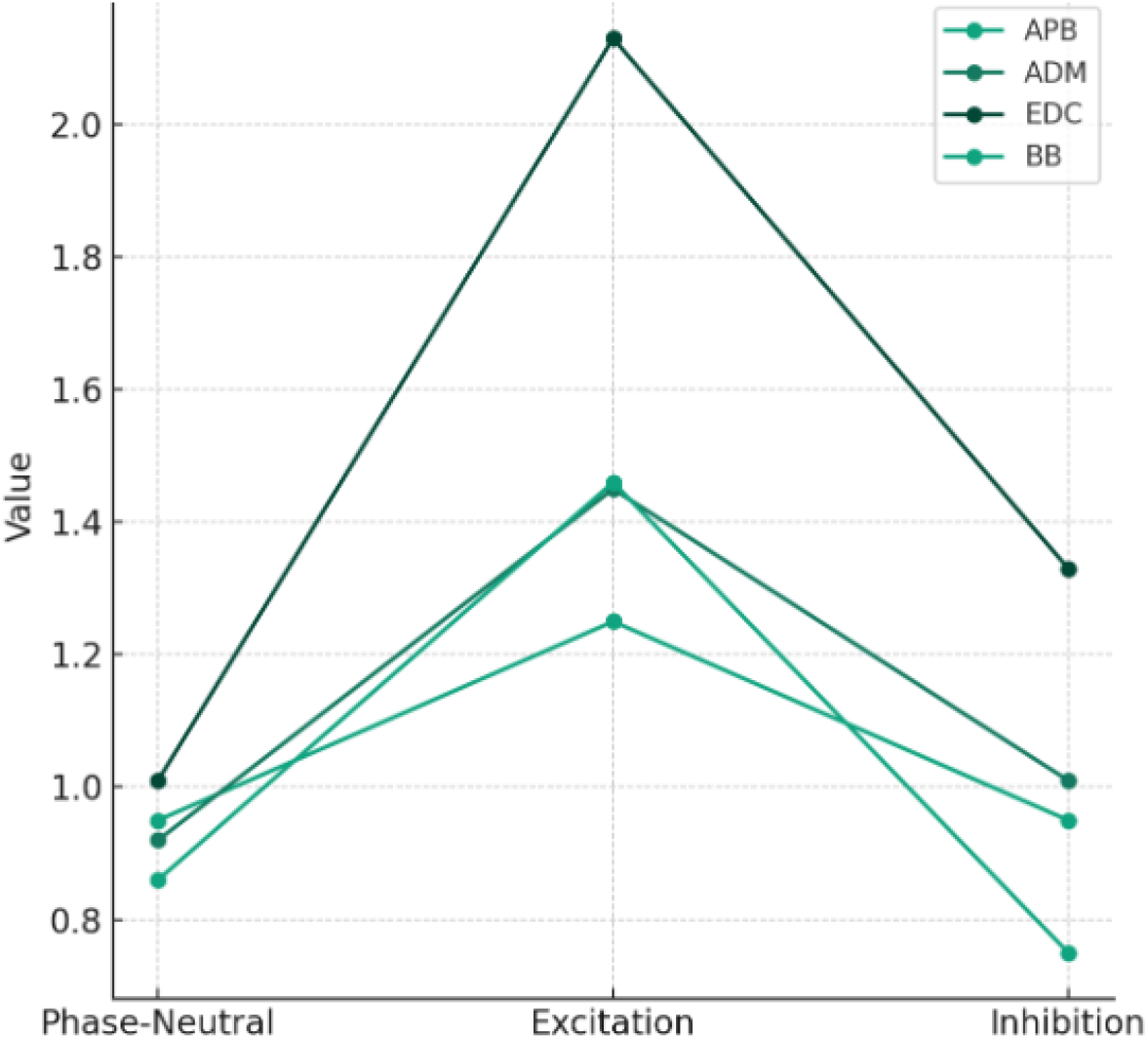
The MCR areas for four different muscles at random, 0°, and 180° phase of sensorimotor rhythm

Although more research is needed it is already possible to observe mechanistically interpretable effects of phase dependency in the spatial maps obtained with the real-time experiment. When introduced into routine clinical practice the demonstrated real-time state-dependent mapping will provide more stable and detailed maps with fewer stimulation pulses than it is currently possible with the phase agnostic approach or using post-hoc SMR phase locking analysis.

## 4 Discussion

Future neuroscientific studies will heavily rely on the tools furnishing an interactive contact with a nervous tissue, either invasive or non-invasive with a small segment of neural tissue, the entire brain or a functional system. To showcase this novel technology in action we developed its embodiment in the form of HarPULL, Hardware Powered Ultra Low Latency oscillatory brain state tracking system and used it to control real-time transcranial magnetic stimulation. HarPULL technology provides its user with robust, reliable and accurate phase tracking with the help of the state-space estimation techniques implemented on board of an EEG device controlled by a truly real-time operational system. HarPULL translates brain’s rhythmic activity parameters such as instantaneous phase and amplitude into a control signal to trigger TMS pulse within less than 2 ms latency from the brain event.

In order to isolate the target rhythmic component a filtering operation is employed. The real-time use (when no future data is available) of traditional static filters causes significant delays (on the order of several hundreds of milliseconds) which hinders brain-machine interaction and results in the feedback signal (e.g. TMS pulse) being delayed by several periods of the target rhythm. The development of advanced solutions for isolating narrow-band components [51] and tracking their state appears to be an important direction towards an interactive contact with neuronal tissue.

The pioneering PHASTIMATE approach [65, 66, 68] solves this problem by building an autoregressive model to forecast the narrow-band filtered (and therefore lagged) time series by the amount of filter’s group delay. This approach has already enabled several innovative studies but its performance critically depends on the forecasting accuracy of the AR model.

HarPULL’s phase tracking algorithm is based on the state-space model estimation by means of the Kalman filter as an optimal approach to identify the analytic signal corresponding to the target rhythmic activity. It was shown earlier that *white* Kalman filter, a simplest form of the KF, used to identify brain rhythm’s model as a frequency modulated process [62] outperforms previously state-of-the-art PHASTIMATE technique in phase tracking accuracy, incurred lag and robustness with respect to the phase resets and central frequency or bandwidth changes [63].

Algorithmic delays are not the only source of the total latency incurred by the modern PC-controlled setups. Data transfer from an EEG device to a PC and the use of a PC running under fundamentally non-real time operating system results in additional variable lags which we aimed to avoid. Described in equations 12 - 14 steady state mode allows for a very compact and low computational cost implementation on board of a system with limited computational resources such as the readily commercially available and cleared for medical purposes NVX-52 EEG device used in this study.

Extending *white* KF approach by including a realistic model of the 1*/f* brain noise, see equations 19, further improves phase tracking accuracy. With simulated data when the ground-truth is known *pink* KF outperformed both *white* KF and PHASTIMATE which emphasizes the importance of not only realistically modelling the target process but also matching closely the dynamic properties of the concurrent additive noise.

During the validation on real data using a publicly available dataset [67] we noticed that the mean error and circular standard deviation increased as compared to the simulated data even for the high SNR segments (SNR *>* 3.0). One of the reasons here is associated with the difficulties in defining the ground-truth phase value in real data as described in details in [67]. In this work we used instantaneous phase time series extracted from the data by means of several non-causal filters as suggested in [67]. Even in this scenario *pink* KF derived instantaneous phase estimates appeared to align better with non-causally derived ground truth as compared to the AR-model based forecasts, see Figure 8.

Most critically, both KFs appear to be not only capable of accurate phase tracking but also introduce minimal delays in the phase estimates, see [63] and our results in Figures 7.a and 8.a. These properties of the KF and its stability reported in [48] made it a method of choice for our HarPULL system capable of robust and zero latency real-time tracking of brain’s rhythm phase. This is achieved by implementing the main processing and generating the corresponding trigger signal on board of a real-time signal processing platform of the NVX-52 amplifier powered by the TMS320VC5509A signal processor designed for fixed latency operation. Modest computational resources were sufficient to accommodate steady state *white* Kalman filter within the reprogrammed core of this commercially available and clinically approved regular EEG machine.

In our system the parameters of the real-time signal processing algorithm are first identified based on a short “training” data and then loaded into the amplifier together with the target phase value. When the instantaneous phase value is reached a TMS pulse is triggered and processing of the EEG time series is suspended for a short time to avoid saturation and a long transient.

We validated our HarPULL setup using phantom experiment, see Figure 10, where the ground truth was exactly known. We have also conducted experiments with a healthy volunteer. The experiments demonstrated the technical feasibility of truly real-time brain rhythm phase-triggered TMS controlled solely by the EEG amplifier and the software running on its operational system.

Finally for the first time in a completely real-time mode and using a PC-free setup we obtained the data to generate muscle cortical representation (MCR) maps. We then visualized the obtained brain-state contingent measurements using the *TMSmap* software [38] (https://tmsmap.ru/), see Figure 12. These maps simply reflect mean MEP amplitudes measured at the target muscle when the stimulation is applied to the prespecified grid of cortical locations. The use of the real-time state dependent approach allowed us to reduce the number of TMS pulses and tailor each TMS pulse to the desired target phase value corresponding to either excitatory or inhibitory states of the sensorimotor system. As expected the MCR maps appeared to critically depend on the SMR phase. On average, for all muscles we observed lower amplitude maps in the inhibitory condition as compared to either state agnostic or the excitation state corresponding to 180°. The MCR map obtained during the excitation state, see Figure 12.b, shows better delineation between the APB and the ADM muscles as compared to either phase agnostic stimulation Figure 12.a or stimulation during the inhibitory phase of the SMR.

Brain rhythms are characterised by a well pronounced transient nature and occur in bursts. From the signal processing point of view rhythm’s phase can only be defined when the rhythm is present, i.e. during its bursts. This fact was recently demonstrated in the context of differentiation between the excitation and inhibition of the sensorimotor system using the SMR’s phase [39]. The SMR’s phase appeared to matter only during the segments with high SNR of the rhythmic activity. HarPULL’s state space tracking identifies both real and imaginary parts of the analytic signal corresponding to the target rhythmic component, see equation 4. The samples of analytic signal furnish a direct access to the rhythm’s instantaneous power which allows to further increase the accuracy of delineation between the excitation and inhibition states. One possible caveat could be the fact that the rhythm’s envelop appears to be delayed by at least 50-100 ms [51] which means that the phase specific stimulation would be delayed with respect to the very onset of the SMR’s burst and will be postponed to its second period since its inception. More advanced algorithms such as those described in [48] are required to further reduce the envelope latency estimation currently achievable with dynamical model based state-space approaches. These solutions, likely to be based on neural networks, will require significant computational resources available onboard of the EEG devices.

Among the limitations of our study is the experimental real-time research made on just two healthy volunteers. The authors have no doubt that more studies are needed, both in healthy population and with patients suffering from neurological and psychiatric diseases. The very question of directional changes in MEPs depending on the phase of the underlying rhythm in these populations requires serious investigation and our system (HarPULL) facilitates such studies.

The proposed solution is likely to further the development of approaches aimed at establishing an interactive contact with the nervous tissue which will find multiple uses in both clinical practice and research. The TMS based mapping allows for non-invasive mapping of motor cortex and visualisation of muscle cortical representations. On the basis of these results it appears possible to derive a range of diagnostic and prognostic biomarkers of both central nervous system damage and the recovery potential. TMS stimulation is used for preoperative mapping of speech and motor cortex. Both conceptually and on the basis of our very preliminary observation we may hope that the use of state-dependent TMS will furnish a more reliable separation of closely located representations of different muscles, will reduce the number of false negative responses, will improve reproducibility and reduce the time required for this procedure.

Also, the state dependent TMS technology is likely to be instrumental in measuring cortical inhibition and facilitation. The disbalance between these two fundamental characteristics of the nervous system appears to be associated with a range of neurological conditions such as multiple sclerosis, ALS, motor cortex hyperexcitability, Parkinson’s decease, etc. Assessment of well established measures of the inhibition-facilitation balance such as short and long interval intracortical inhibition (SICI LICI), and short-interval interhemispheric inhibition (SIHI) is likely to become more robust when performed in the brain-state aware mode which is facilitated by the proposed HarPULL technology.

In our work we combined HarPULL with TMS as a convenient platform to rapidly demonstrate the effects of the instantaneous context dependent interaction with the nervous tissue. At the same time the uses of HarPULL as a technology for real-time tracking of nervous tissue’s oscillatory state are versatile. Coupled with tACS our solution holds significant importance due to its potential to precisely modulate neural oscillations and synchronize or desynchronize brain activity by tuning the phase of the alternating current with respect to that of the brain’s native oscillatory activity. With this tool researches will be able to explore dynamic neural underpinnings of various cognitive functions such as attention, memory, and perception. Our solution combined with tACS will also become instrumental as a basis for therapeutic applications in a range of neurological and psychiatric disorders including Parkinson’s and epilepsy when it is necessary to disrupt the pathologicall activity of the native neural populations.

Finally, HarPULL combined with neurofeedback and stimulus presentation devices using conventional sensory pathways (audio, tactile, video) will be useful in cognitive studies exploring the influence of rapidly evolving brain activity on the efficacy of incoming information processing. Combined with electrophysiological signals reflecting activity of autonomous nervous system HarPULL will be instrumental in studying complex brain-body interactions, a novel and rapidly developing branch of modern neuroscience.

## Data Availability

All data produced in the present study are available upon reasonable request to the authors

## 5 Acknowledgements

The research leading to these results has received funding from the Basic Research Program at the National Research University Higher School of Economics.

